# Genetic evidence for efficacy of targeting IL-2, IL-6, and TYK2 signaling in prevention of type 1 diabetes: A Mendelian randomization study

**DOI:** 10.1101/2023.12.15.23300016

**Authors:** Tea E Heikkilä, Emilia K Kaiser, Jake Lin, Dipender Gill, Jaakko J Koskenniemi, Ville Karhunen

## Abstract

**Background:** Type 1 diabetes is an autoimmune disease, which leads to insulin dependence. We investigated genetic evidence to support the repurposing of seven drugs, already licensed or in clinical phases of development, for prevention of type 1 diabetes.

**Methods:** We obtained genome-wide association study (GWAS) summary statistics for the risk of type 1 diabetes, whole-blood gene expression, and serum protein levels, and investigated genetic polymorphisms near seven potential drug target genes. We used colocalization to examine whether the same genetic variants that are associated with type 1 diabetes risk were also associated with the relevant drug target genetic proxies, and Mendelian randomization to evaluate the direction and magnitude of the associations. Furthermore, we performed Mendelian randomization analysis restricted to functional variants within the drug target genes.

**Findings:** Colocalization revealed that the blood interleukin (IL)-2 receptor subunit alpha (*IL2RA*) and IL-6 receptor (*IL6R*) gene expression levels within the corresponding genes shared the same causal variant with type 1 diabetes liability (posterior probabilities 100% and 96.3%, respectively). Odds ratios (OR) of type 1 diabetes per 1-SD increase in the genetically proxied gene expression of *IL2RA* and *IL6R* were 0.22 (95% confidence interval [CI] 0.17-0.27) and 1.98 (95% CI 1.48-2.65), respectively. Using missense variants, genetically proxied tyrosine kinase 2 (*TYK2*) expression levels were associated with type 1 diabetes risk (OR 0.61, 95% CI [0.54-0.70]).

**Interpretation:** Our findings support the targeting of IL-2, IL-6R and TYK2 signaling in prevention of type 1 diabetes. Further studies *should* assess the optimal window to intervene to prevent type 1 diabetes.

**Funding:** Kyllikki ja Uolevi Lehikoisen säätiö, Foundation for Pediatric Research, Finnish Cultural Foundation, JDRF International; The University of Oulu & The Research Council of Finland Profi 326291; European Union’s Horizon 2020 research and innovation program under grant agreement no. 848158 (EarlyCause).

**Research in context:** *Evidence before this study:* We searched PubMed for GWAS, Mendelian randomization, colocalization, and other studies for genetic evidence of efficacy of targeting IL2RA, IL2RB, IL6R, IL6ST, IL23A, TYK2, JAK2, and JAK3 signaling in prevention of type 1 diabetes. We searched studies with combinations of “IL2RA”, “IL2RB”, “IL6R”, “IL6ST”, “IL23A”, “JAK2”, “JAK3”, “TYK2”, “colocalization”, "mendelian randomization", or “GWAS” and "Diabetes Mellitus, Type 1"[Mesh]. A recent large GWAS by Robertson et al. observed five conditionally independent genome-wide significant single-nucleotide polymorphisms (SNPs) associated with the risk of type 1 diabetes in intergenic and intronic areas near *IL2RA*. Another large recent GWAS by Chiou et al. found six independent SNPs associated with the risk of type 1 diabetes near *IL2RA*. A few smaller candidate gene studies also reported associations between the risk of type 1 diabetes and eight other loci near IL2RA. Epigenetic studies reported that a few type 1 diabetes risk variants near *IL2RA* are associated with changes in methylation of DNA near promoters of *IL2RA* in white blood cells and B-cells. Both above mentioned GWAS found associations between two missense mutations in *TYK2* gene (rs12720356(A>C), rs34536443(G>C)) and the risk of type 1 diabetes. The same variants protected against type 1 diabetes and some other autoimmune diseases in a Finnish FINNGEN biobank study. Smaller candidate gene studies found that the rs2304256 A/A genotype was protective against type 1 diabetes in a Southern Brazilian and in a European population. In a study of Japanese population, similar association was not seen, possibly due to differences in allele frequencies and incidence of type 1 diabetes between populations. In the same study, TYK2 promoter haplotype with genetic polymorphisms in promoter region and exon 1 decreased the promoter activity and increased the risk for type 1 diabetes. The minor 358Ala allele of the IL6R SNP rs2228145 (A>C) was found to be associated with a reduced risk of developing T1D. In addition to loci near or at IL2RA and TYK2 loci, the GWAS by Robertson et al. reported an association between rs2229238 near *IL6R* and the risk of type 1 diabetes. Furthermore, a candidate gene study found that the GG haplotypes of variants rs11171806 and rs2066808 (in strong LD) near *IL23A* was protective against T1D. We found no studies in which colocalization between the loci near the reviewed genes and type 1 diabetes was reported nor estimates from Mendelian randomization on the possible causal relationship. However, Robertson et al. studied the genetic evidence for therapeutic potential of their GWAS hits using a priority index algorithm, which combined information from GWAS of type 1 diabetes, expression quantitative loci in immune cells, evidence from chromatin conformation in immune cells, as well as protein-protein interactions and genomic annotations. Among the reported GWAS hits, priority index ranked IL2RA as the top target, followed by TYK2 (the 3rd target), JAK2 (11th), IL23 (25^th^), JAK3 (35^th^), and IL6R (50^th^).

*Added value of this study:* We found that the risk of type 1 diabetes and whole-blood gene expression of *IL2RA* and *IL6R* colocalized to rs61839660 near *IL2RA* and rs10908839 near *IL6R*, respectively. Using these loci as instruments of *IL2RA* and *IL6R* expression, Mendelian randomization indicated that whole blood *IL2RA* expression is associated with decreased risk of type 1 diabetes (OR 0.70 per 1 SD increase) whereas IL6R expression is associated with increased risk (OR 1.079 per 1 SD increase). Furthermore, when a missense variant rs2304256 in *TYK2* was used as an instrument for *TYK2* expression, Mendelian randomization indicated that an SD increase in *TYK2* expression was predicted to decrease the risk of type 1 diabetes (OR 0.61).

*Implications of all the available evidence:* We found genetic evidence supporting the efficacy of targeting IL-2, IL-6 and TYK2 signalling in prevention of type 1 diabetes. Further research is needed to address when would be the most optimal time to intervene in the pathogenesis of type 1 diabetes.

## Introduction

Type 1 diabetes is an autoimmune disease characterized by the loss of beta cell function. Despite advances in continuous glucose monitoring and insulin administration, managing type 1 diabetes remains a significant burden and few patients reach glycemic targets^1^. Several drugs have shown potential in delaying the loss in beta cell function in newly diagnosed cases^2^, and teplizumab, a monoclonal anti-CD3 antibody, even delayed the onset of the clinical disease by an average of two years^3^. However, no current therapy can completely halt the disease progression, and therefore it is crucial to identify new efficacious drug targets.

Drug targets backed by genetic evidence have higher success rates in clinical development^4^. Genome-wide association studies (GWAS) enable the discovery of genomic regions strongly associated with the disease of interest. These associations can be considered as evidence for the involvement of the corresponding proteins in the disease pathogenesis, implying these proteins are potential drug targets for the disease. The evidence for a likely drug target can be further investigated via colocalization and Mendelian randomization. Colocalization can be utilized to study whether the same causal variant is shared between the drug target and the disease liability, or if the loci of risk allele and an allele influencing the drug target are distinct^5^. Furthermore, Mendelian randomization can be used to assess how much the genetic variability in drug target levels affects the risk of a disease in the population^6^.

To inform the prioritization of targets for prevention of type 1 diabetes, we aimed to investigate the genetic evidence for efficacy of 12 drug targets in prevention of type 1 diabetes. These targets were selected because a prior GWAS reported that they are associated with the risk of type 1 diabetes and drugs that target them are already licensed for other indications than type 1 diabetes or they have progressed to clinical development (Supplementary Table 1).

## Methods

### Study design

We selected the drug targets based on a previous GWAS of type 1 diabetes by Robertson et al.^7^ (Table 1; Supplementary Figure 1). Utilizing a priority index, the study ranked the drugs based on four factors including 1) existence of genetic variant(s) close to the potential target, 2) chromatin accessibility, 3) gene expression data in relevant cell types, and 4) protein-protein interactions^8^. We focused on 12 proteins (IL2RA, IL2RB, IL2RG, IL6R, IL6ST, IL12B, IL23A, IFNAR2, JAK1, JAK2, and JAK3, TYK2) which have already been targeted in clinical trials for autoimmune diseases and some of which have been licensed for other indications than type 1 diabetes (Supplementary Table 1)^7^. Among these targets, eight target genes (IL2RA, IL2RB, IL6R, IL6ST, IL23A, JAK2, JAK3, TYK2) had a locus associated with the risk of type 1 diabetes (p<1·10^-5^) within the distance of 1 million base pairs from the target gene (see Supplementary Figure 2). IL23A was excluded from the analyses because the pQTL data were unavailable, leaving seven targets for the subsequent analyses. Genomic regions under investigation are listed in Table 1.

**Table 1.** Genomic regions under investigation.

| Gene | Chromosome | Start position<sup>1</sup> | End position<sup>1</sup> | Variants available<sup>2</sup> |
| --- | --- | --- | --- | --- |
| *IL2RA* | 10 | 6,010,689 | 6,062,367 | 9,683 |
| *IL2RB* | 22 | 37,118,666 | 37,175,118 | 5,489 |
| *IL23A* | 12 | 56,338,884 | 56,340,410 | No pQTL data available |
| *IL6R* | 1 | 154,405,193 | 154,469,450 | 4,274 |
| *IL6ST* | 5 | 55,935,095 | 55,995,022 | 12,024 |
| *JAK2* | 9 | 4,984,390 | 5,129,948 | 7,939 |
| *JAK3* | 19 | 17,824,780 | 17,848,071 | 7,142 |
| *TYK2* | 19 | 10,350,533 | 10,380,608 | 6,403 |

<sup>1</sup>Using genome build hg38. <sup>2</sup>Within $\pm 1$ million base pairs from the gene.

All primary studies that generated the GWAS summary statistics used in our analysis have undergone institutional board review and received ethical approval^9–11^.

### Participants

We obtained the genome-wide association study (GWAS) summary statistics for type 1 diabetes, whole-blood gene expression (expression quantitative trait loci, eQTL) and serum protein levels (protein quantitative trait loci, pQTL) (Supplementary Table 2). pQTL data were used for loci in the vicinity of *IL6ST* since it codes a soluble protein, whereas eQTL data were used for loci near the other drug target genes since they code intracellular or membrane-bound proteins. We included loci within 1 Mb from the seven potential drug target genes (Table 1). We obtained the data on type 1 diabetes risk variants from a subsequent GWAS of 18,942 cases and 501,638 controls of European ancestry from 9 cohorts^9^. We obtained pQTL data for IL6ST from a GWAS of 35,559 Icelanders and eQTL data from GWAS of 31,684 individuals from 37 eQTLGen Consortium cohorts, most individuals being European^10,11^. We have summarized the study population details of the utilized GWAS as well as methods of ascertainment of cases of type 1 diabetes, measurement of whole-blood gene expression, and serum protein levels in Supplementary note 1.

### Colocalization

We conducted colocalization analysis to assess whether the genetic associations for type 1 diabetes risk near the seven drug target genes align with those for the whole-blood gene expression or serum protein levels of these targets. We performed a colocalization analysis using a ’coloc’ package in R^5^. This method utilizes Bayesian principles to assess the relationship between two traits. It considers all variants within a specific genetic locus and evaluates the following hypotheses, assuming a maximum of one causal variant per trait:

H0: There is no association with either trait, implying no specific causal variants. H1: There is an association with the exposure trait only.
H2: There is an association with the outcome trait only.
H3: There are associations with both exposure and outcome traits, two independent SNPs i.e., distinct casual variants.
H4: There are associations with both exposure and outcome traits, one shared causal variant.

A high posterior probability for H4 implies a shared causal variant for the two traits. A substantial posterior probability for H3 suggests the two traits are influenced by distinct causal variants linked to each trait. We used the default prior probabilities of 10^-4^, 10^-4^ and 10^-5^ for a variant being associated with the exposure trait, the outcome trait, and both traits, respectively.

### Mendelian randomization

To evaluate the direction and magnitude of the causal effects, we performed Mendelian randomization for those variants that colocalized between drug target levels and the risk of type 1 diabetes (posterior probability for H4 >0.8). Mendelian randomization utilizes genetic variants to investigate the relationship between an exposure (e.g. drug target levels) and an outcome (e.g. type 1 diabetes risk) for causality. Under Mendel’s law of assortment, genetic variants are accepted to be independent of other genetic alleles and can be used as valid instrumental variables to estimate the causal effect of the exposure on the outcome if the three assumptions are fulfilled:

1: the genetic variant is associated with the exposure,
2: the genetic variant is associated with the outcome only through the exposure, and 3: the genetic variant is not associated with any confounders^6^.

Advantages of Mendelian randomization include limited susceptibility to reverse causation and confounding by external factors that influence both exposure and the outcome. Linkage disequilibrium (LD) may confound Mendelian randomization if genetic variants are not shared between the exposure and outcome, but they reside in the same genomic area. However, we investigated this possibility in the prior colocalization step.

### Examination of functional variants near drug target genes

To further examine the potential causality of the putative drug targets on type 1 diabetes risk, we sought for functional missense variants in coding area of these 7 drug target genes from Phenoscanner^12^ that were associated with the protein/expression levels of the target at p<10^-5^. We used a more lenient threshold than the genome-wide significance of p<5e-8 since we only focused on cis-variants. The threshold can be interpreted as a Bonferroni-corrected threshold for 5,000 independent variants. These variants were then individually used as instruments in Mendelian randomization to test for causality of the targets on the risk of type 1 diabetes.

### Statistical analysis

All analyses were done with R version 4.2 (R Foundation for Statistical Computing, Vienna, Austria) using packages ‘coloc’, ‘hyprcoloc’, and ‘TwoSampleMR’^5,13,14^. Effect sizes for change in the risk of type 1 diabetes is reported per change in standard deviation of serum protein or mRNA levels, and the genetic variants were not weighted in any of the analyses. Beta and SE values for whole-blood RNA levels were calculated using formulae 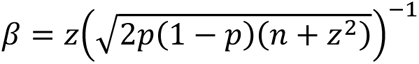 and SE = 2p(1 − p)(n + z^2^)^−1^, where p = minor allele frequency, n=sample size, and z = z-score. Mendelian randomization estimates are reported as Wald estimates. We only used variants that were available in the GWAS summary statistics for both exposure and outcome traits within each genomic locus. All the LD r^2^ values reported in this study were obtained from the European population of the 1000 Genomes project using the package ‘ieugwasr’. The study protocol was not preregistered for this study. The code used to generate our results is available at https://github.com/jkoskenniemi/T1DSCREEN.

## Results

### The same genetic variants affect type 1 diabetes risk and whole-blood IL2RA and IL6R gene expression

We found evidence for a shared causal variant between the risk of type 1 diabetes and whole-blood *IL2RA* gene expression (rs61839660, posterior probability 100%, Figure 1) and type 1 diabetes and whole-blood *IL6R* gene expression (rs10908839, posterior probability 96.5%, Figure 2). The variant rs10908839 is in strong LD (r^2^ = 0.76) with the previously identified lead SNP rs2229238 associated with the risk of type 1 diabetes in the study by Robertson et al^7^. No evidence for colocalization was observed between drug target levels and the risk of type 1 diabetes near the other target genes (Table 2, Supplementary Figures 3-7).

**Figure 1.**
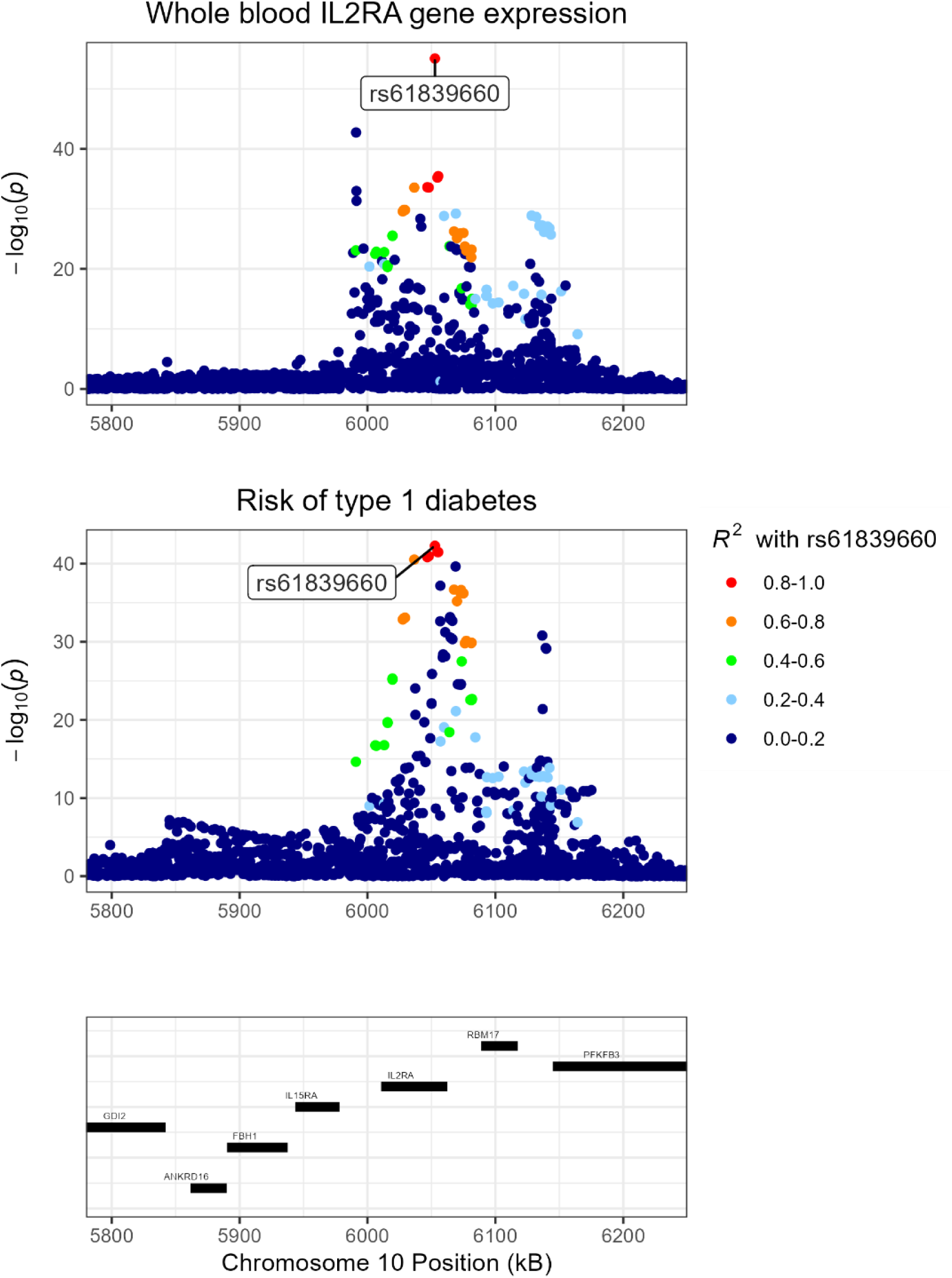
Regional Manhattan plot of *IL2RA* gene expression and risk of type 1 diabetes near *IL2RA* gene. The colours indicate the linkage disequilibrium (LD) *r2* value (based on 1000Genomes European reference) with rs61839660, the most likely shared causal variant identified in colocalization.

**Figure 2.**
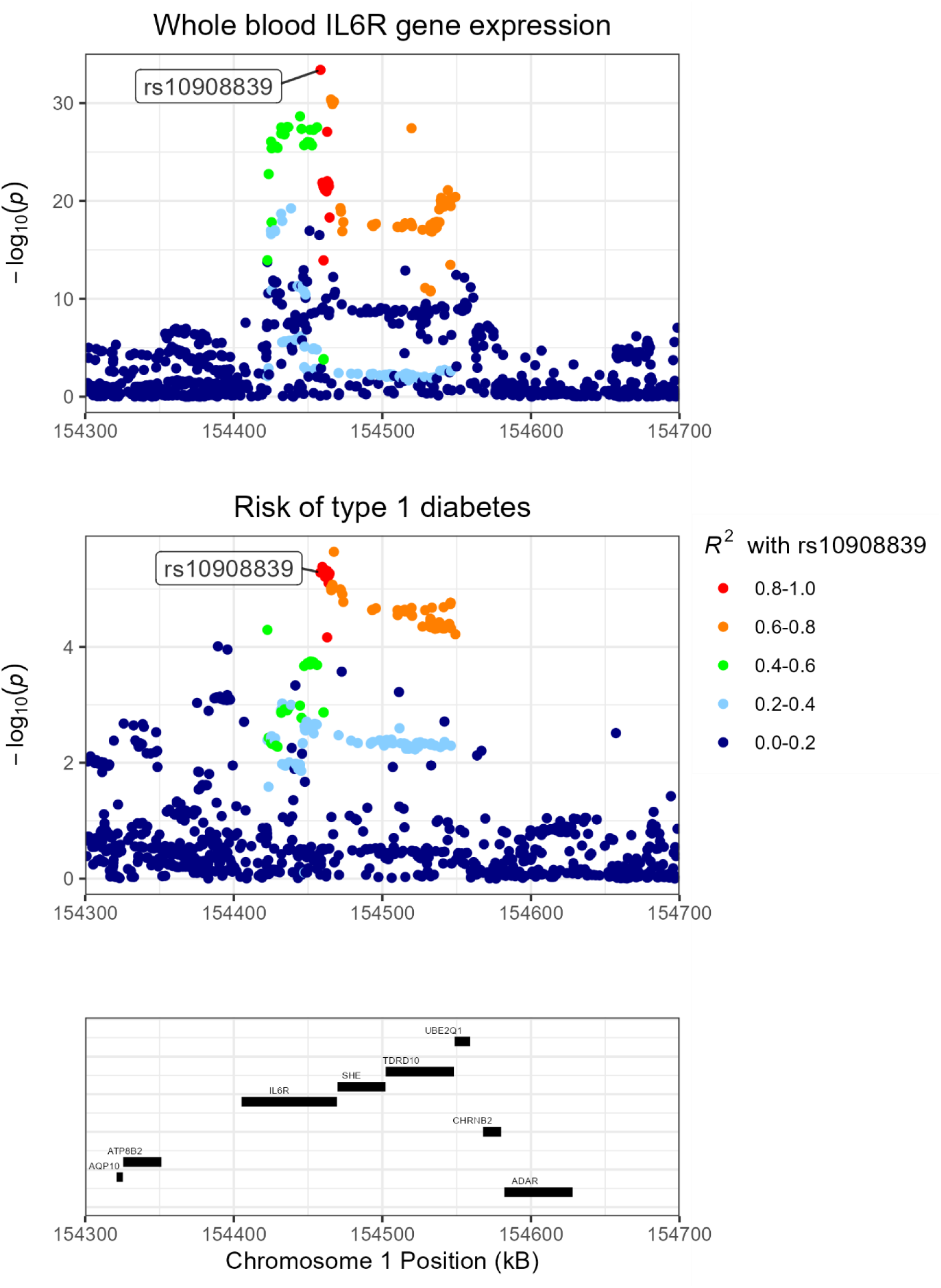
Regional Manhattan plot of *IL6R* gene expression and risk of type 1 diabetes near *IL6R* gene. The colours indicate the linkage disequilibrium (LD) *r2* value (based on 1000Genomes European reference) with rs10908839, the most likely shared causal variant identified in colocalization.

**Table 2.** Results of colocalization between drug target (eQTL/pQTL) and risk of type 1 diabetes.

| Gene | Trait | H0 | H1 | H2 | H3 | H4 |
| --- | --- | --- | --- | --- | --- | --- |
| *IL2RA* | eQTL | 0.0 % | 0.0 % | 0.0 % | 0.0 % | **100.0 %** |
| *IL2RB* | eQTL | 0.0 % | 0.0 % | 0.0 % | **99.7 %** | 0.3 % |
| *IL6R* | eQTL | 0.0 % | 0.5 % | 0.0 % | 2.9 % | **96.5 %** |
| *IL6ST* | pQTL | 0.0 % | 9.3 % | 0.0 % | **90.5 %** | 0.2 % |
| *JAK2* | eQTL | 0.0 % | 0.0 % | 0.0 % | **100.0 %** | 0.0 % |
| *JAK3* | eQTL | 0.0 % | 1.2 % | 0.3 % | **98.5 %** | 0.0 % |
| *TYK2* | eQTL | 0.0 % | 0.0 % | 0.0 % | **86.6 %** | 13.4 % |

*eQTL = expression quantitative trait loci, pQTL = protein quantitative trait loci. H0: no association with either trait, H1: an association with the exposure trait only, H2: an association with the outcome trait only, H3: associations with both exposure and outcome traits, two independent SNPs i.e., distinct casual variants. H4: associations with both exposure and outcome traits, one shared causal variant.*

### Genetically proxied variability in IL2RA and IL6R gene expression is associated with the risk of type 1 diabetes

We investigated the direction and magnitude of the expected change in the risk of type 1 diabetes if IL2RA and IL6R were targeted using rs61839660 near *IL2RA* and rs10908839 near *IL6R* as genetic instruments of *IL2RA* and *IL6R* expression in Mendelian randomization. Variant rs61839660 was associated with *IL2RA* gene expression (β=0.23, se=0.015, p=8.8·10^-56^) and variant rs10908839 with *IL6R* gene expression (β=0.11, se=0.0092, p=4.1·10^-34^) in whole-blood, and both variants were associated with the risk of type 1 diabetes (odds ratio [OR] 0.70, 95% confidence interval [95% CI] 0.66-0.74, p=5.3·10^-43^ and OR 1.079, 95% CI 1.04-1.12, p=5.2·10^-6^, respectively). Mendelian randomization revealed an odds ratio (OR) of 0.22 (95% CI 0.17-0.27) on type 1 diabetes risk (p=5.3 · 10^-43^) per SD increase in genetically proxied *IL2RA* expression and OR of 1.98 (95% CI 1.48-2.65) per SD increase in genetically proxied *IL6R* expression (p=5.2 · 10^-6^).

### Functional variants in coding area of IL2RA, IL6R, and TYK2 are associated with the risk of type 1 diabetes

We found 22 missense mutations in the examined drug target genes with GWAS summary data on type 1 diabetes risk (Table 3). Most of the mutations had an allele frequency of <0.01 with notable exceptions of rs2228145 (MAF **=** 0.39) in *IL6R* as well as rs12720356 (MAF = 0.09) and rs2304256 (MAF=0.28) in *TYK2.* Summary statistics for both drug target level and type 1 diabetes risk were available for nine variants. One independent variant at LD r^2^<0.2 within each locus (rs41316003 in *JAK2*, rs2304256 in *TYK2*, and rs141500365 *IL6ST*) was associated with the relevant drug target (protein or gene expression levels at p<10^-5^). When using these variants as instruments in Mendelian randomization, the genetically proxied *TYK2* gene expression was associated with the risk of type 1 when using missense variants in *IL6ST* (OR 0.98, 95% CI 0.78-1.20), or *JAK2* (OR 0.74, 95% CI 0.46-1.20) as shown in Supplementary Table 3. Supplementary Table 4 shows the LD matrix between the functional variants in coding area of *TYK2* as well as the lead SNPs associated with *TYK2* expression and the risk of type 1 diabetes in the vicinity of *TYK2* gene.

**Table 3.** Functional variants in the examined 7 drug targets.

| | SNP | MAF | EA | NEA | $\beta_{T1D}$ | SE<sub>T1D</sub> | p<sub>T1D</sub> | $\beta_{QTL}$ | p<sub>QTL</sub> |
| --- | --- | --- | --- | --- | --- | --- | --- | --- | --- |
| *IL2RA* | rs72650666 | 0.004 | A | G | 0.18 | 0.15 | 0.22 |  |  |
| *IL2RB* | rs149508414 | 0.006 | T | C | -0.27 | 0.11 | 0.02 |  |  |
|  | rs143857582 | 0.002 | T | C | -0.094 | 0.19 | 0.62 |  |  |
| *IL6R* | rs2228145 | 0.39 | C | A | -0.044 | 0.014 | 0.002 |  |  |
| *IL6ST* | rs34417936 | 0.02 | T | C | -0.11 | 0.049 | 0.03 | -0.11 | $5.4 \cdot 10^{-4}$ |
| | rs61748224 | 0.003 | T | C | 0.058 | 0.14 | 0.68 | -0.34 | $7.8 \cdot 10^{-4}$ |
| | rs141500365 | 0.002 | A | G | 0.044 | 0.22 | 0.84 | -1.9 | $4.7 \cdot 10^{-138}$ |
|  | rs61755738 | 0.004 | C | A | -0.015 | 0.11 | 0.89 | -0.008 | 0.91 |
| *JAK2* | rs41316003 | 0.007 | A | G | -0.11 | 0.089 | 0.22 | 0.36 | $1.3 \cdot 10^{-7}$ |
|  | rs143103233 | 0.001 | G | C | -0.065 | 0.23 | 0.77 |  |  |
|  | rs150221602 | 0.001 | C | G | -0.022 | 0.25 | 0.93 |  |  |
| *JAK3* | rs55778349 | 0.008 | C | G | 0.02 | 0.094 | 0.83 |  |  |
| *TYK2* | rs34536443 | 0.04 | C | G | -0.39 | 0.039 | $1.5 \cdot 10^{-23}$ | | |
| | rs2304256<sup>a</sup> | 0.28 | A | C | -0.12 | 0.016 | $1.4 \cdot 10^{-14}$ | 0.25 | $6.4 \cdot 10^{-168}$ |
| | rs12720356<sup>b</sup> | 0.09 | C | A | -0.15 | 0.025 | $2.4 \cdot 10^{-9}$ | 0.17 | $7.0 \cdot 10^{-22}$ |
| | rs35018800 | 0.008 | A | G | -0.4 | 0.091 | $1.4 \cdot 10^{-5}$ | 0.33 | $6.3 \cdot 10^{-4}$ |
|  | rs55762744 | 0.01 | T | C | -0.2 | 0.076 | 0.007 | -0.021 | 0.71 |
|  | rs144960992 | 0.001 | G | A | 0.37 | 0.23 | 0.11 |  |  |
|  | rs55886939 | 0.002 | C | T | -0.23 | 0.19 | 0.21 |  |  |
|  | rs55882956 | 0.002 | A | G | -0.23 | 0.21 | 0.27 |  |  |
|  | rs188833543 | 0.003 | T | C | -0.15 | 0.14 | 0.29 |  |  |
|  | rs139954108 | 0.0003 | G | A | -1 | 1.5 | 0.49 |  |  |

SNP = single-nucleotide polymorphism, MAF = minor allele frequency, EA = effect allele, NEA = non-effect allele, $\beta_{T1D}$ = effect size for the risk of type 1 diabetes, SE<sub>T1D</sub> = standard error for the effect size for the risk of type 1 diabetes, p<sub>T1D</sub> = p-value for the risk of type 1 diabetes, $\beta_{QTL}$ = beta coefficient for effect size for gene or protein expression, p<sub>QTL</sub> = p-value for gene or protein expression. The values $\beta_{QTL}$ for and p<sub>QTL</sub> are provided only if the corresponding data were available.
<sup>a</sup>rs2304256 is in very high LD with the lead eQTL rs34725611 ( $r^2=1.00$ ) and in moderate LD with the lead T1D risk variant rs144309607 ( $r^2=0.10$ ).
<sup>b</sup>rs12720356 is in LD with the other TYK2 eQTL missense variant rs2304256 ( $r^2=0.29$ ).

## Discussion

Leveraging data from large-scale GWAS, we investigated the genetic evidence for efficacy of seven candidate drugs in prevention of type 1 diabetes. Using colocalization and Mendelian randomization, we found genetic evidence to support the role of IL-2 and IL-6 signaling in the pathogenesis of type 1 diabetes. In addition, the investigation of functional missense variants suggested that TYK2 signaling is involved in the etiology of type 1 diabetes.

Our evidence for the protective effect of *IL2RA* expression on the risk of type 1 diabetes supports the role of regulatory T-cells as a natural protection against type 1 diabetes. Low, but sufficient level of IL-2 is crucial for the survival and function of regulatory T-cells^15^. On the other hand, high concentrations of IL-2 during acute infections predominantly boost immune response by stimulating effector T-cells and natural killers (NK), while the effect of IL-2 on regulatory T-cells maintains a level of tolerance towards self-peptides. These characteristics of immune system are achieved by the differences in affinity to IL-2 between the high-affinity heterotrimeric receptor in regulatory T-cells, which consists of α-, β-, and γ-chains encoded by *IL2RA, IL2RB*, and *IL2RG* genes, respectively, and intermediate-affinity heterodimeric receptor (consisting of only β- and γ-chains) in other cells that express only IL2RB and IL2RG^16^. Thus, our finding supports the rationale of prevention trials of type 1 diabetes with low dose IL-2^17^, which expands the population of regulatory T-cells without excessively amplifying effector T-cell or NK cell populations^18^.

In accordance with our finding of involvement of whole-blood IL6R signaling in pathogenesis of type 1 diabetes, a previous observational study reported higher C-reactive protein (CRP) levels, a read-out of IL-6 signaling^19^, in children with type 1 diabetes compared to healthy individuals^20^. The immune system increases IL-6 production rapidly in response to infections and tissue damage, which promotes various acute phase responses such as CRP production^19^. Previous experimental studies support our finding of the role of IL-6 signaling in pathogenesis of type 1 diabetes. IL-6 inhibits the development and function of regulatory T-cells (Treg) and promotes the development of pathogenic T-helper 17 (Th17) cells^21^. Abnormalities in IL-6 signaling lead to Treg/Th17 imbalance, which is a causative factor in autoimmune diseases such as rheumatoid arthritis^21^, and this is also suggested to occur in type 1 diabetes^22^. IL-6 signaling occurs as classic signaling through a cell membrane-bound IL6R, trans-signaling through soluble circulating IL6R and soluble gp130 (encoded by *IL6ST*), and trans-presentation by dendritic cells, in which IL6 is presented to T-cells via dendritic cell membrane-bound IL6R^23^.

In contrast to our findings, tocilizumab, a monoclonal antibody against IL6R, did not significantly affect the decline in residual beta cell function in individuals with newly diagnosed type 1 diabetes in a randomized, placebo-controlled, double-blind clinical trial^24^. This discrepancy may be partially explained by the timing of the intervention. Genetic polymorphisms typically exert life-long influence on risk of diseases and affect every stage of the disease’s development, whereas the intervention in the study by Greenbaum et al. took place after the diagnosis of type 1 diabetes, when a sharp fall in beta cell function had already taken place^25^.

While whole-blood TYK2 expression and the risk of type 1 diabetes did not colocalize, this absence of evidence may reflect violations to the one-causal-variant assumption in the ‘coloc’ method. Indeed, the missense mutation rs2304256 identified in *TYK2* gene is in very high LD (r^2^ =1.00) with the lead eQTL rs34725611. Moreover, rs2304256 is only in moderate LD (r^2^ =0.10) with rs144309607, the lead variant on T1D liability in colocalization, suggesting two independent signals. Of note, a known missense variant rs34536443 is not available in the eQTL data and therefore could not be used in colocalization. Despite the one-causal-variant assumption, ‘coloc’ is relatively robust to multiple causal variants, and colocalization methods allowing for multiple causal variants are highly sensitive to LD misspecifications in the reference panel^6^. Therefore, in the absence of an accurate LD reference, we proceeded with the ‘coloc’ method for our colocalization while acknowledging its limitations.

Overall, our *TYK2* findings are consistent with previous studies suggesting that TYK2 signaling is associated with the risk of type 1 diabetes^26^. Promising results have been found in clinical trials targeting TYK2 in autoimmune diseases supporting the potential of drug repurposing in type 1 diabetes^27^. Other reasons for the colocalization discordance could be that TYK2 is not activated until IFN alpha binding to IFNAR1 and TYK2 RNA expressions are known to have low tissue and cell type specificity^28,29^.

It is important to note that Mendelian randomization estimates are only valid if the instrumental variable associations (relevance, independence, exclusion restriction) are met. The strong associations between the assessed genetic polymorphisms and the studied exposures suggested that the genetic instruments studied were relevant for the studied exposures. C*is*-Mendelian randomization studies are by design less prone to violations of independence and exclusion restriction assumptions, as genetic variants typically exert the strongest influence on nearby genes and therefore most effects are secondary to reading of the nearby genes. Furthermore, our colocalization results suggest that association between whole-blood *IL2RA* and *IL6R* gene expression and the risk of type 1 diabetes are unlikely to be caused by LD with a genetic variant that primarily influences the reading of other genes in the vicinity of *IL2RA* or *IL6R* loci.

Some aspects of generalizability of our results are also worth mentioning. The original GWAS studies primarily included individuals of European descent, which limits the spectrum of rare variants and the generalizability of our findings to other ancestries. Furthermore, our Mendelian randomization estimates represent the influence of small changes in *IL2RA*, *IL6R* and *TYK2* gene expression during the entire life course before the diagnosis of type 1 diabetes. Therefore, these effect sizes cannot be directly extrapolated to clinical trials in which the doses are larger and exposures shorter and possibly outside a key sensitive window for disease development. Thus, natural history studies and clinical prevention trials should pinpoint the optimal stage of pathogenesis when to interfere with IL-2, IL-6, or TYK2 signaling to prevent type 1 diabetes.

In conclusion, our results provide genetic evidence that IL-2 and IL-6 signaling, and regulatory T-cells are associated with type 1 diabetes. Our findings encourage clinical trials that investigate the efficacy of drugs such as tocilizumab (IL6R antagonist) and low dose aldesleukin (IL-2 analogue) in prevention of type 1 diabetes.

## Data Availability

The code used to generate our results is available at https://github.com/jkoskenniemi/T1DSCREEN.

## Acknowledgements

The authors wish to acknowledge CSC – IT Center for Science, Finland, for computational resources and original studies for summary statistics.

## Declaration of generative AI and AI-assisted technologies in the writing process

During the preparation of this work the authors used ChatGPT version 4.0 to proof-read and to improve the readability of the manuscript. After using this tool/service, the authors reviewed and edited the content as needed and take full responsibility for the content of the publication.

## Declaration of interests

Authors declare no competing interests.

## Supplementary Note 1

We obtained genome-wiwde association study (GWAS) summary statistics for type 1 diabetes, whole-blood gene expression (expression quantitative trait loci, eQTL) and serum protein levels (protein quantitative trait loci, pQTL). The serum protein levels were measured with 4,907 aptamers in 35,559 Icelanders using the SomaScan multiplex aptamer assay (version 4), including 4,907 relevant aptamers, which target 4,719 proteins. Plasma samples were collected from 40,004 Icelanders between 2000 and 2019. A total of 52% of the participants were from the Icelandic Cancer Project (newly diagnosed cancer patients and their relatives and controls selected from the National Registry) and 48% were from genetic programs at deCODE genetics, mainly from the population-based deCODE Health study. In the Icelandic Cancer Project, participants were patients with newly diagnosed cancer and their relatives. The average age of the participants was 55 years and 57% of the participants were women^1^.

The data for gene expression was obtained from 31,684 individuals through 37 eQTLGen Consortium cohorts. 25,482 of the samples were from whole blood and 6,202 were peripheral blood mononuclear cell samples. The majority of samples were of European ancestry and from population-based cohorts. Gene expression levels of samples were measured by Illumina, Affymetrix, Affymetrix Hu-Ex version 1.0 ST expression arrays and RNA-seq. In a large-scale meta-analysis, cis-eQTL was identified for 16,987 genes, trans-eQTL for 6,298 genes and eQTS effects for 2,568 genes. In cis-eQTL, gene expression levels are affected by a proximal (<1Mb) SNP and in trans-eQTL by a distal (>5Mb) SNP or a SNP on another chromosome^2^.

The GWAS data for the risk of type 1 diabetes is from nine cohorts including 18,942 individuals with type 1 diabetes and 501,638 control individuals of European ancestry. The type 1 diabetes case cohorts were combined to the control cohorts based on genotyping array (Affymetrix, Illumina Infinium, Illumina Omni, and Immunochip) and ancestry (Ireland, UK or USA). For the GENIE-UK cohort, no matched country of origin was found for the control cohort so individuals of British ancestry from the University of Michigan Health and Retirement study (HRS) were used as controls. For the UK Biobank cohort and non-UK Biobank cohorts, non-European ancestry samples were removed. For control cohorts, phenotype files were used to remove individuals with type 2 diabetes or autoimmune diseases. ICD10 was used in the UK Biobank cohort to identify 1,445 type 1 diabetes cases (diagnosis of type 1 diabetes and insulin treatment within a year of diagnosis, no type 2 diabetes diagnosis). The controls had no diagnosis of type 1 or type 2 diabetes or autoimmune disease^3^.

---

**Supplementary Table 1.**
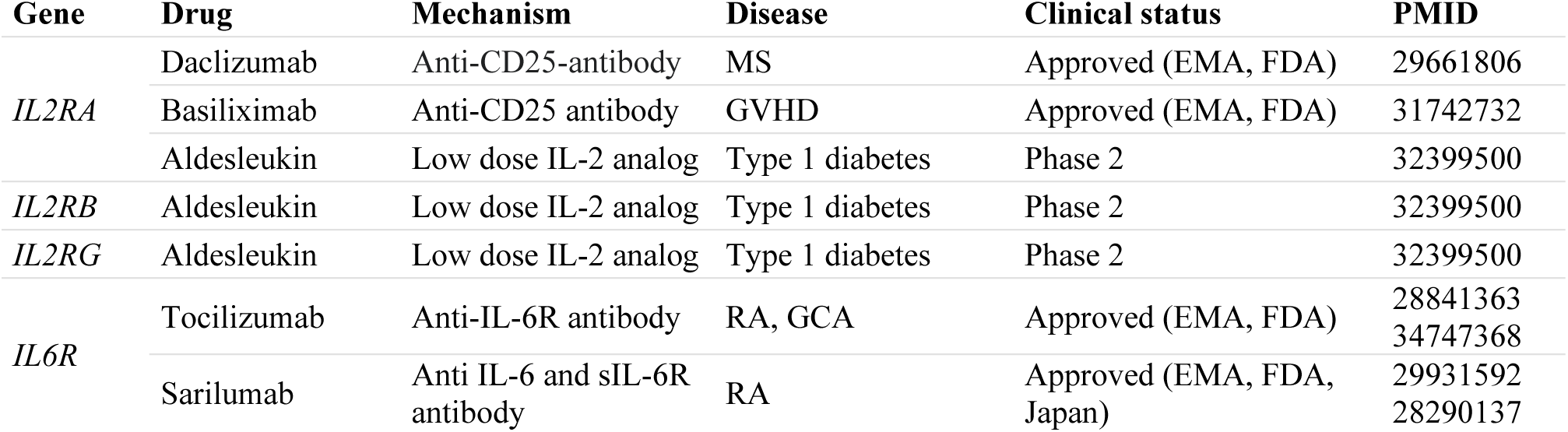
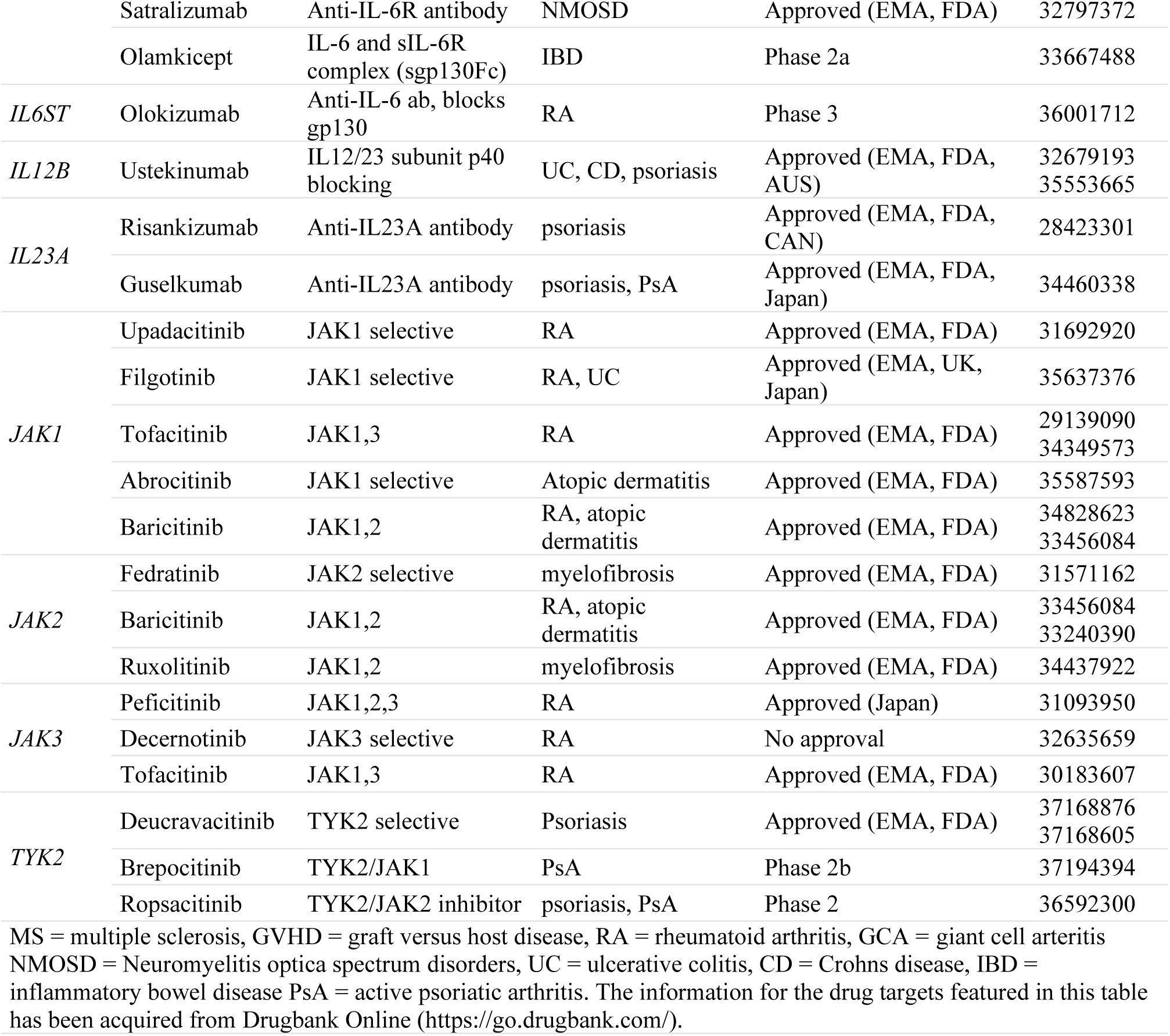
Selected drug targets.

**Supplementary Table 2.** Source studies of GWAS summary statistics.

| Trait | Study | DOI | N |
| --- | --- | --- | --- |
| Risk of type 1 diabetes | Chiou et al. 2021 | 10.1038/s41586-021-03552-w | 18,942 cases, 501,638 controls |
| Serum protein levels (except IL23A) | Ferkingstad et al. 2021 | 10.1038/s41588-021-00978-w | 35,559 |
| Whole blood gene expression data | Võsa et al. 2021 | 10.1038/s41588-021-00913-z | 31,684 |

**Supplementary Table 3.** Mendelian randomization effect sizes of four functional variants.

| | SNP | $\beta$ | SE | OR | 95%CI | p |
| --- | --- | --- | --- | --- | --- | --- |
| *IL6ST* | rs141500365 | -0.023 | 0.11 | 0.98 | 0.78-1.20 | 0.84 |
| *JAK2* | rs41316003 | -0.30 | 0.25 | 0.74 | 0.46-1.20 | 0.22 |
| *TYK2* | rs2304256<sup>a</sup> | -0.49 | 0.064 | 0.61 | 0.54-0.69 | $1.4 \cdot 10^{-14}$ |
| | rs12720356<sup>b</sup> | -0.87 | 0.15 | 0.42 | 0.31-0.56 | $2.4 \cdot 10^{-9}$ |

$\beta$ = beta coefficient for Mendelian randomization, p = p-value for Mendelian randomization, SE = standard error for Mendelian randomization, OR = odds ratio for Mendelian randomization, 95% CI = confidence interval for Mendelian randomization, <sup>a</sup>rs2304256 is in very high LD with the lead eQTL rs34725611 ( $r^2=1.00$ ) and in moderate LD with the lead T1D risk variant rs144309607 ( $r^2=0.10$ ), <sup>b</sup>rs12720356 is in LD with the other *TYK2* eQTL missense variant rs2304256 ( $r^2=0.29$ ).

**Supplementary Table 4.** Linkage disequilibrium (LD) matrix (r^2^) between four genetic variants near *TYK2* gene.

|  | rs34536443<sup>a</sup> | rs12720356<sup>b</sup> | rs2304256<sup>c</sup> | rs34725611<sup>d</sup> | rs144309607<sup>e</sup> |
| --- | --- | --- | --- | --- | --- |
| rs34536443<sup>a</sup> | 1.00 | <0.01 | 0.11 | 0.11 | 0.81 |
| rs12720356<sup>b</sup> | <0.01 | 1.00 | 0.29 | 0.29 | <0.01 |
| rs2304256<sup>c</sup> | 0.11 | 0.29 | 1.00 | 1.00 | 0.10 |
| rs34725611<sup>d</sup> | 0.11 | 0.29 | 1.00 | 1.00 | 0.11 |
| rs144309607<sup>e</sup> | 0.81 | <0.01 | 0.10 | 0.11 | 1.00 |

<sup>a</sup>A known missense variant, which has previously reported to be associated with the risk of type 1 diabetes. <sup>b</sup>The second eQTL and type 1 diabetes risk variant among the missense variants in *TYK2* gene. <sup>c</sup>Top eQTL and type 1 diabetes risk variant among the missense variants in *TYK2* gene in this study. <sup>d</sup>The lead eQTL in the vicinity of *TYK2* gene. <sup>e</sup>The lead type 1 diabetes risk variant near *TYK2* gene. LD data based on the 1000 Genomes project European subsample.

**Supplementary Figure 1.**
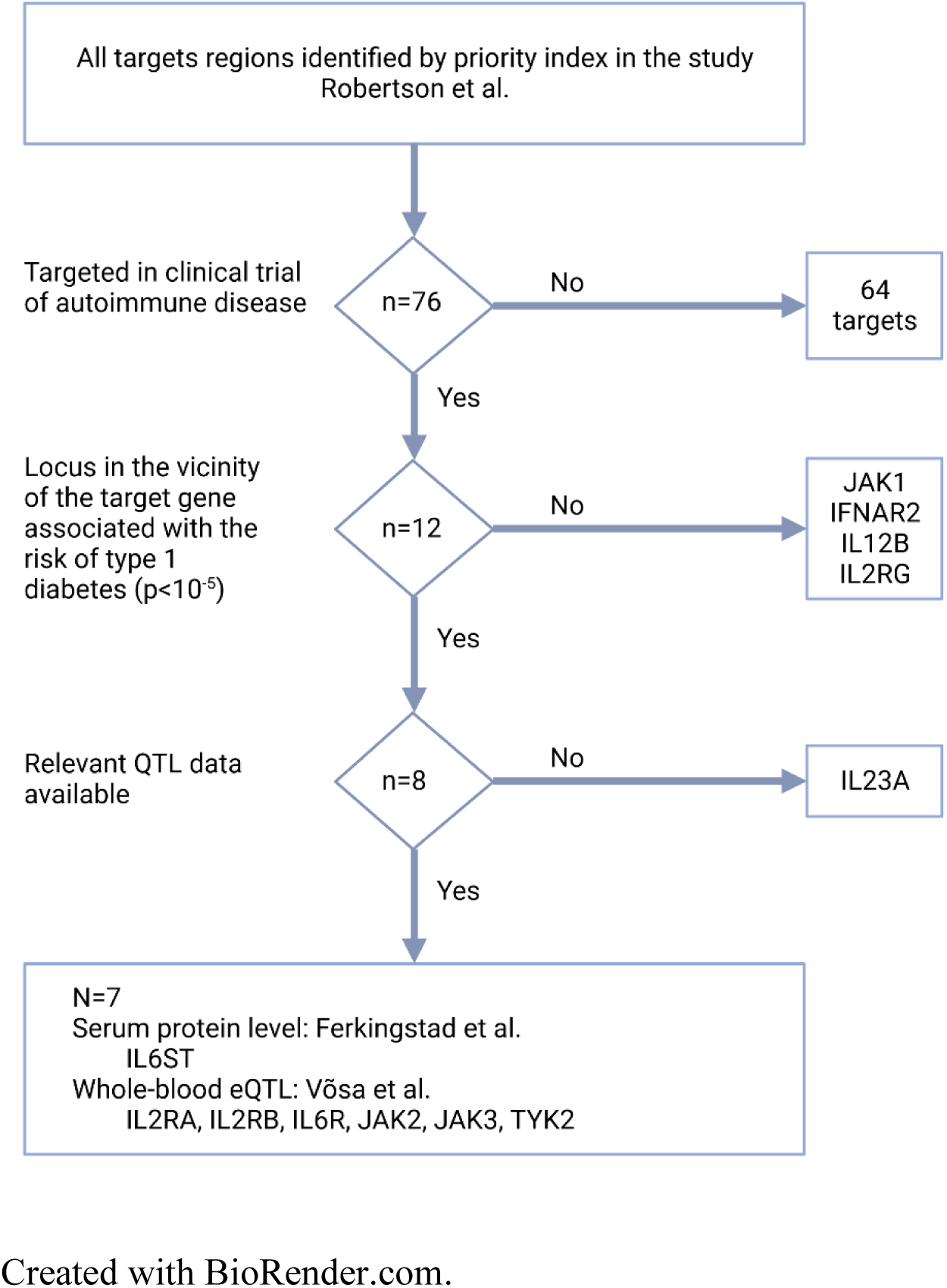
Flow chart of the selection of the targets.

**Supplementary Figure 2.**
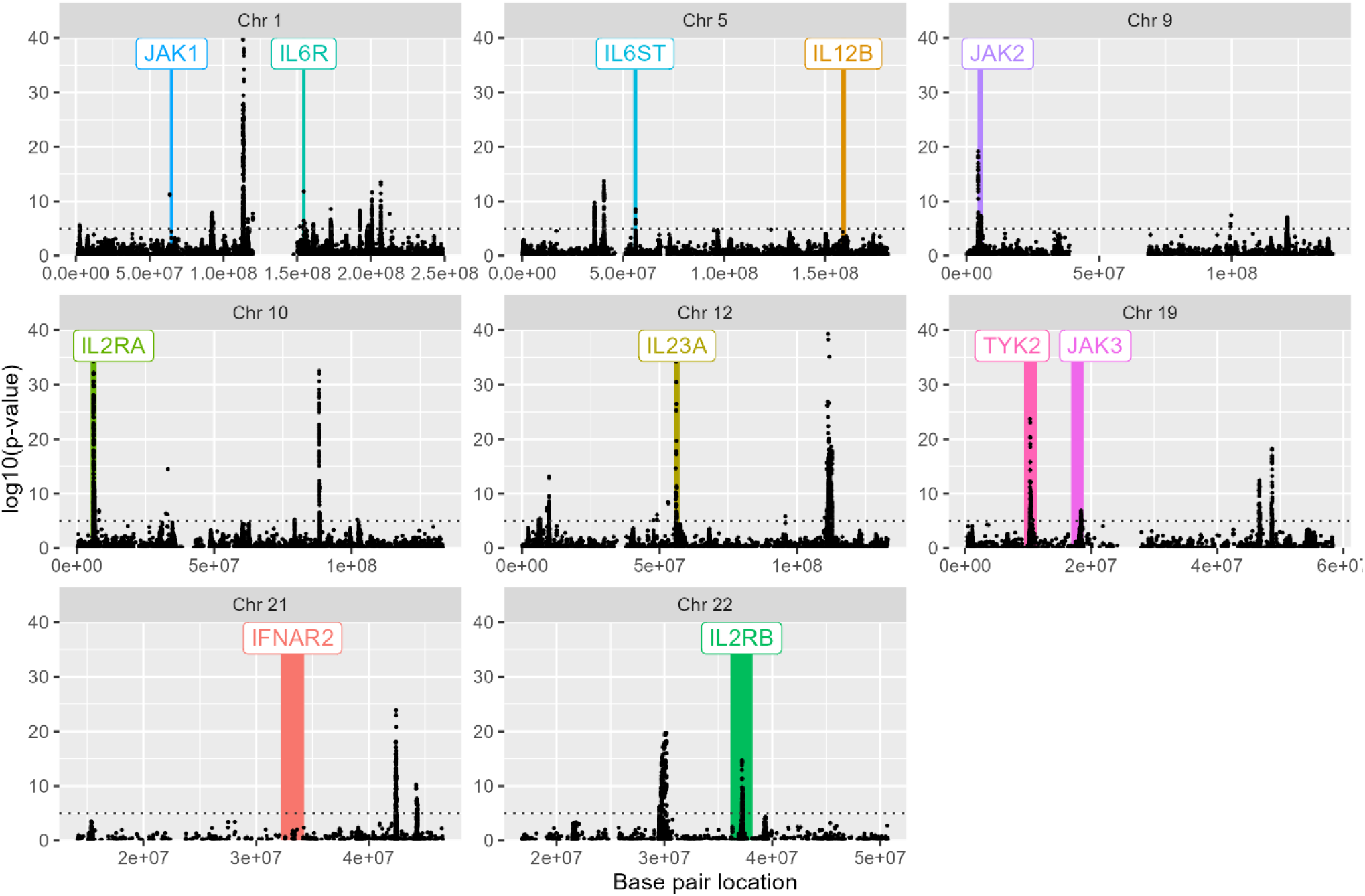
Selected genomic areas of interest in the study Roberston et al. P-values < 10^−40^ are excluded from this figure. Dotted line indicates p=10^−5^. Chr = Chromosome.

**Supplementary Figure 3.**
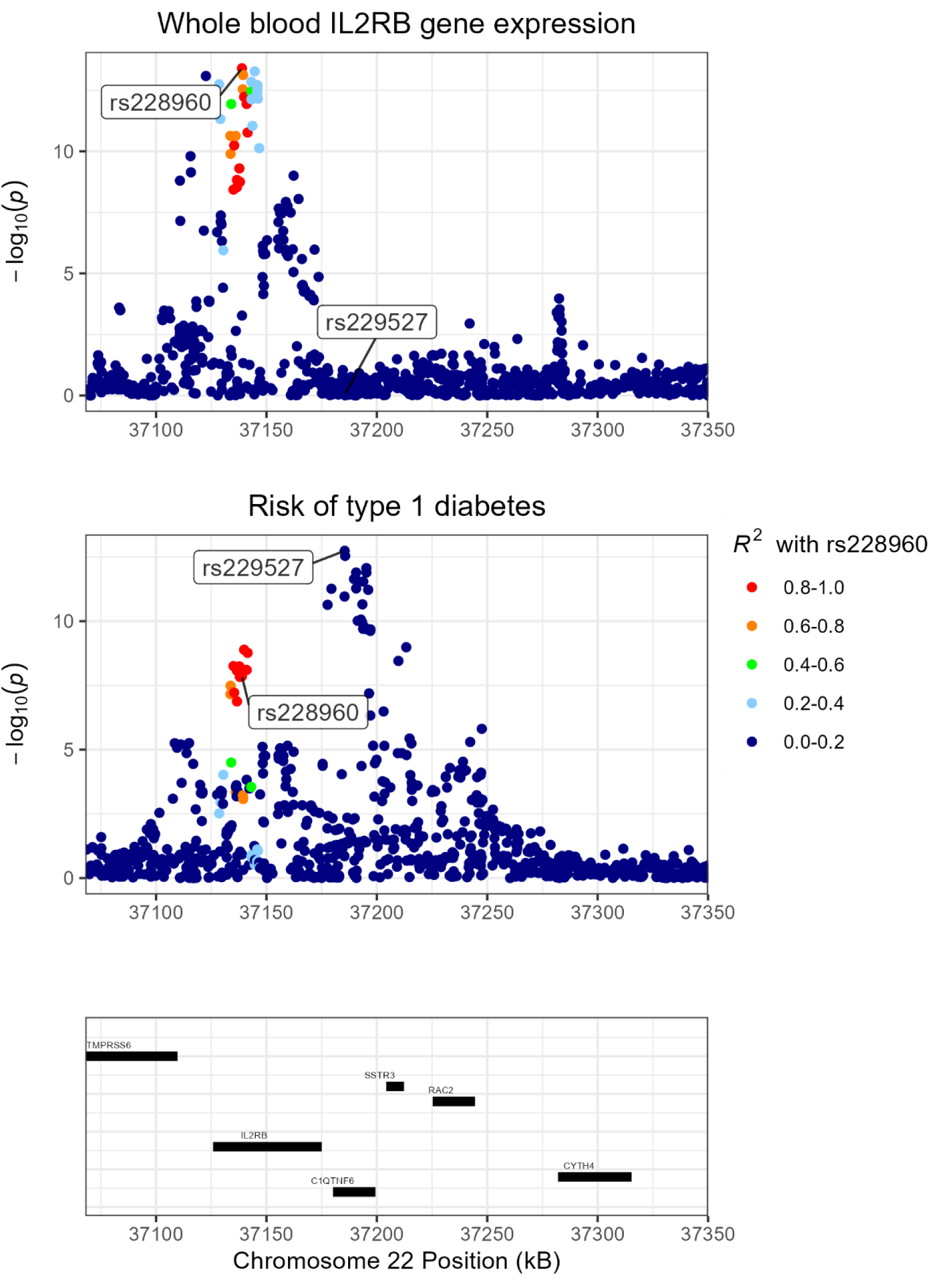

**Supplementary Figure 4.**
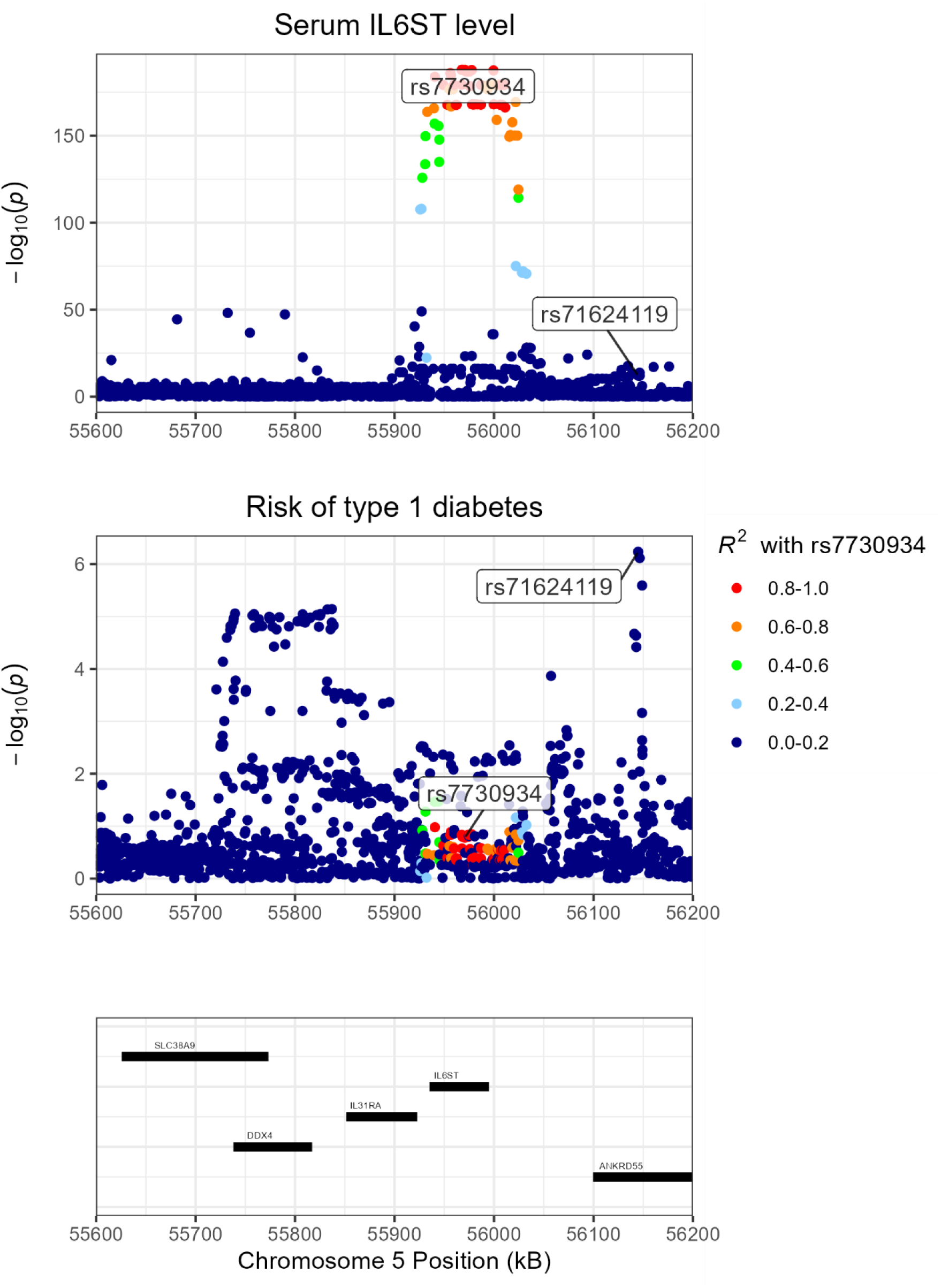

**Supplementary Figure 5.**
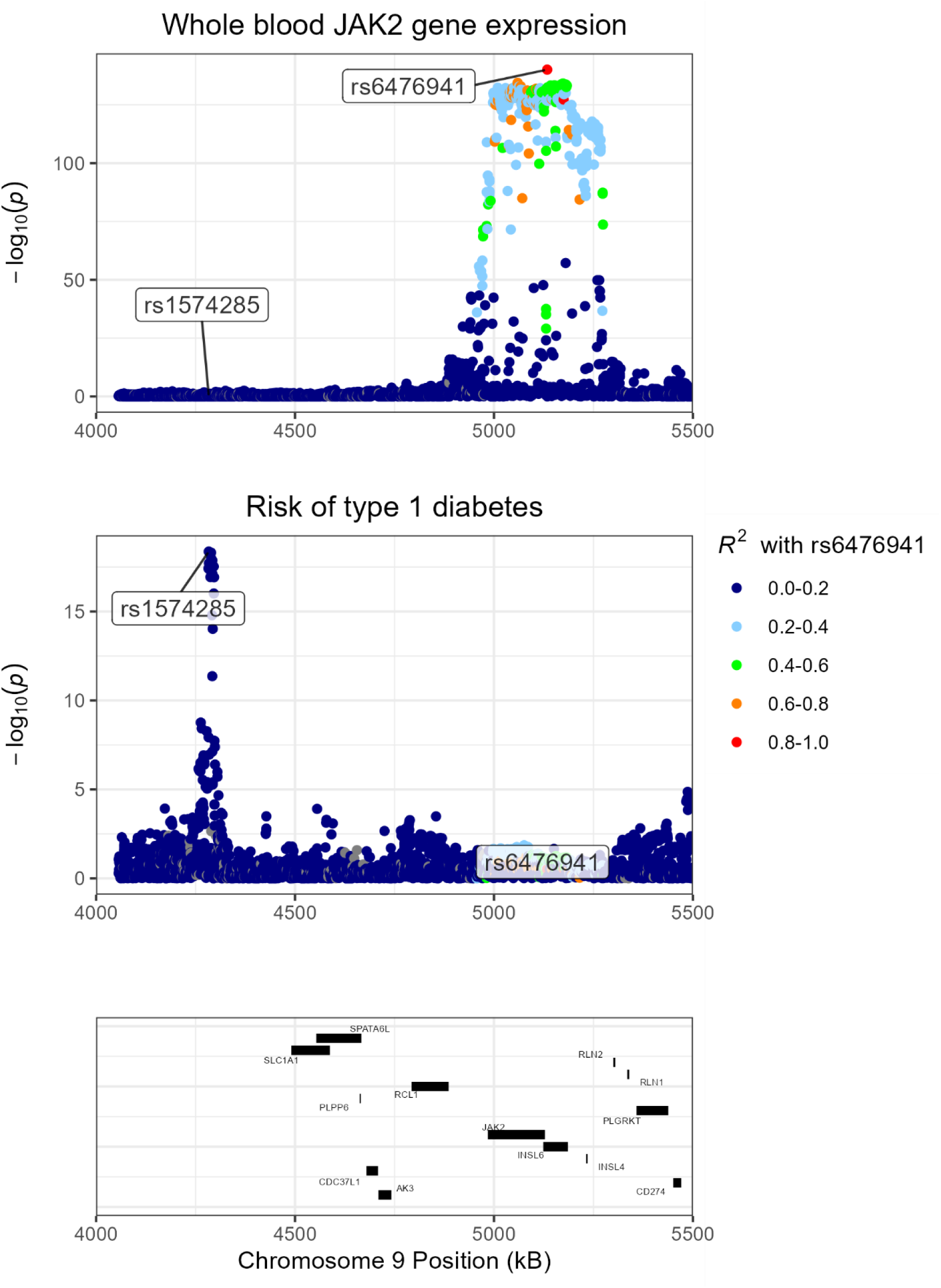

**Supplementary Figure 6.**
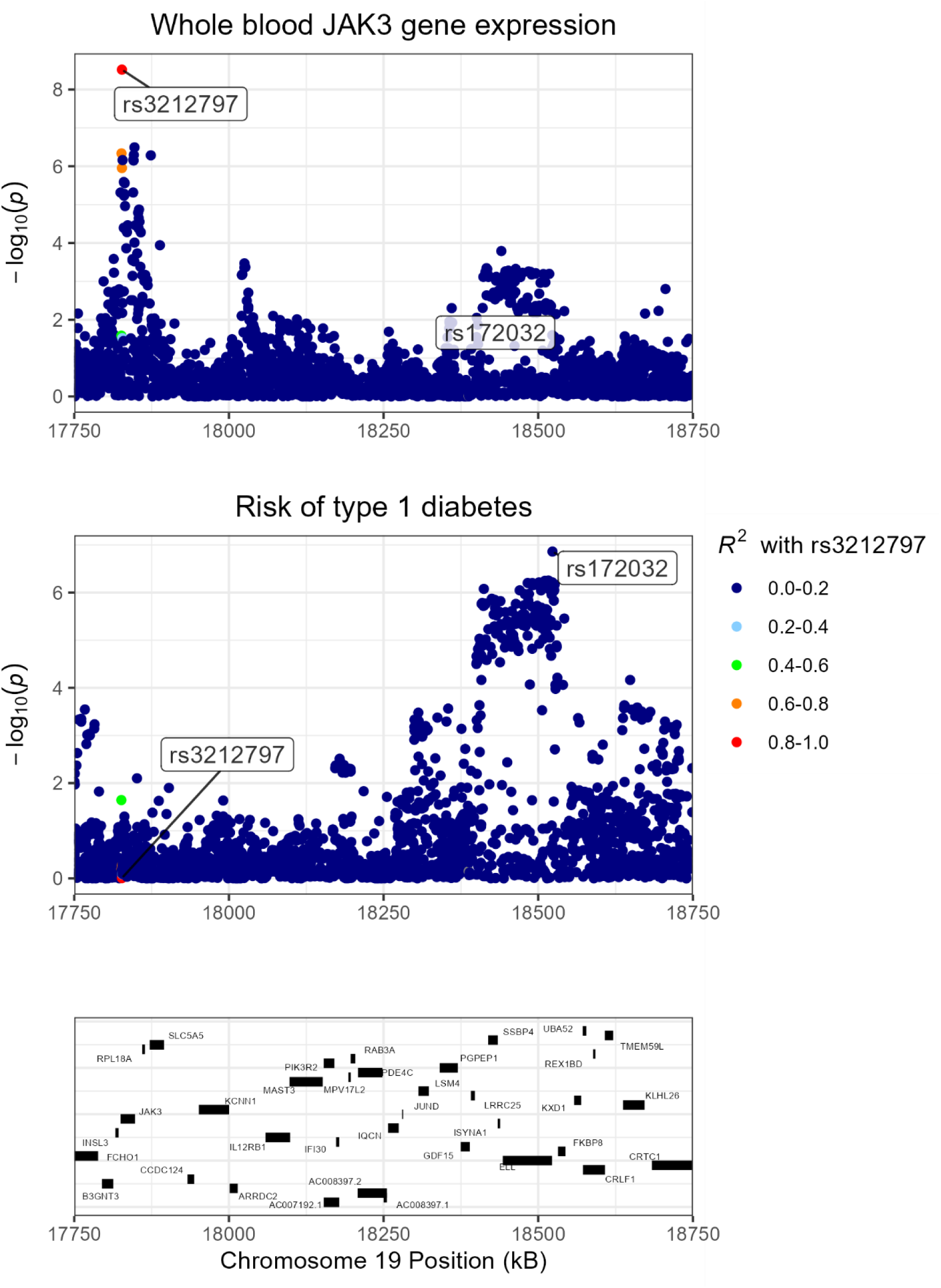

**Supplementary Figure 7.**
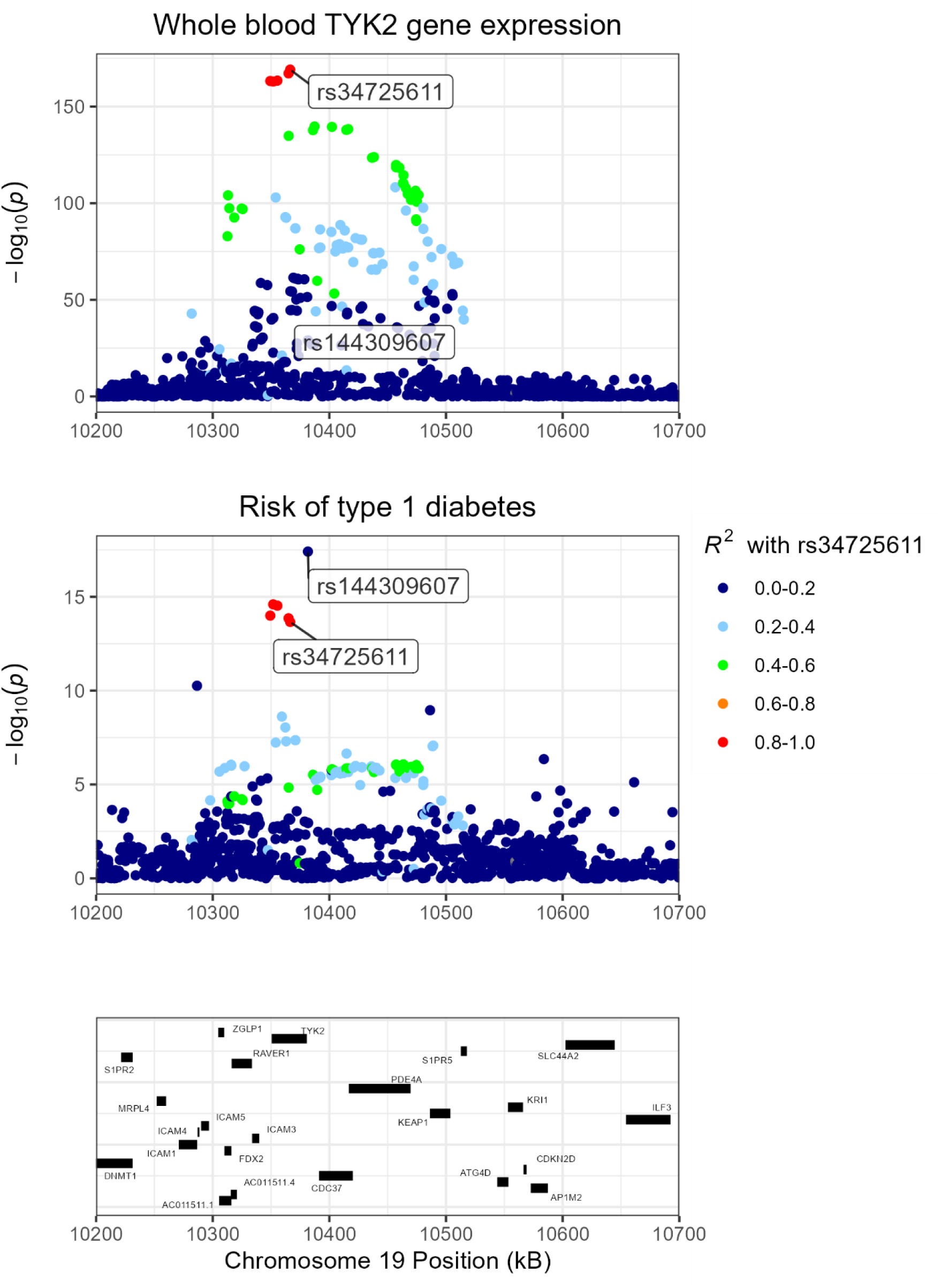

## Notes

### Competing Interest Statement

The authors have declared no competing interest.

### Funding Statement

Kyllikki ja Uolevi Lehikoisen saatio, Foundation for Pediatric Research, Finnish Cultural Foundation, JDRF International; The University of Oulu & The Research Council of Finland Profi 326291; European Union's Horizon 2020 research and innovation program under grant agreement no. 848158 (EarlyCause).

### Author Declarations

We used publicly available summary data from published studies that obtained relevant ethical approval and participant consent. The code used to generate our results is available at https://github.com/jkoskenniemi/T1DSCREEN.

